# An evaluation of assumptions underlying respondent-driven sampling and the social contexts of sexual and gender minority youth participating in HIV clinical trials in the United States

**DOI:** 10.1101/2020.11.02.20222489

**Authors:** AL Wirtz, J Iyer, D Brooks, K Hailey-Fair, N Galai, C Beyrer, D Celentano, R. Arrington-Sanders, PUSH Study Group

## Abstract

**Introduction:** Respondent-driven sampling has been an effective sampling strategy for HIV research in many settings, but has had limited success among some youth in the United States. We evaluated a modified RDS approach for sampling Black and Latinx sexual and gender minority youth (BLSGMY) and evaluates how lived experiences and social contexts of BLSGMY youth may impact traditional RDS assumptions.

**Methods:** RDS was implemented in three cities to engage BLSGMY in HIV prevention or care intervention trials. RDS was modified to include targeted seed recruitment from venues, internet, and health clinics, and provided options for electronic or paper coupons. Qualitative interviews were conducted among a sub-sample of RDS participants to explore their experiences with RDS. Interviews were coded using RDS assumptions as an analytic framework.

**Results:** Between August 2017 and October 2019, 405 participants were enrolled, 1,670 coupons were distributed, with 133 returned, yielding a 0.079 return rate. The maximum recruitment depth was 4 waves among seeds that propagated. Self-reported median network size was 5 (IQR 2-10) and reduced to 3 (IQR 1-5) when asked how many peers were seen in the past 30 days. Qualitative interviews (n=27) revealed that small social networks, peer trust, and targeted referral of peers with certain characteristics challenged network, random recruitment, and reciprocity assumptions of RDS. HIV stigma and research hesitancy were barriers to participation and peer referral.

**Conclusions:** Small social networks and varying relationships with peers among BLSGMY challenge assumptions that underlie traditional RDS. Modified RDS approaches, including those that incorporate social media, may support recruitment for community-based research but may challenge assumptions of reciprocal relationships. Research hesitancy and situational barriers must be addressed in recruitment and study designs.

## Introduction

Respondent driven sampling (RDS) has gained popularity in HIV research over the last two decades as a means to sample populations for whom a sampling frame is unavailable.[1, 2] This has typically included populations who are most affected by the HIV epidemic, such as sexual and gender minoritized populations (SGM) who have sex with men (transgender women and gay and bisexual men). The popularity of RDS is owed to its dual benefits of enabling access to populations who may otherwise be challenging to recruit and the estimation of population prevalence (RDS inference) through the use of weighted estimates.[2-4] Several studies have demonstrated that lengthy referral chains enable recruitment to reach deep into social networks and engage individuals with greater risk behaviors and who may be less connected to services, leading to recent use of RDS for HIV intervention research.[5-7]

Numerous international investigations demonstrate the effectiveness of RDS to engage adult SGM populations in HIV research[8, 9] particularly where same sex relationships or gender identities may be criminalized or stigmatized and communities have subsequently forged strong networks.[10-12] In these contexts peer recruitment effectively builds on social networks among individuals with shared experiences. Recently, however, studies in the US have demonstrated challenges in achieving target sample size and recruitment depth for Black and Latinx SGM populations and SGM youth (SGMY), resulting in modifications of RDS methods to attain study recruitment targets.[13-15]

There are assumptions that are inherent within RDS, which are critical to effective, representative recruitment and inference. These assumptions include: 1) the target population is well networked, 2) peer relationships are reciprocal (undirected network), 3) recruitment within one’s social network is random, and 4) the sample is selected with replacement.[16] The violation of these assumptions on population interferences has been explored in reviews and simulation studies,[17] though less is known about individual participant experiences and behaviors that may challenge these assumptions, particularly how the social context of limited disclosure, stigma, and racism experienced in Black and Latinx SGMY (BLSGMY) may impact RDS assumptions.[18]

The US national strategy, *Ending the HIV Epidemic (EHE)*,[19] is concentrated on the engagement of those most affected by the HIV epidemic – including BLSGMY in HIV surveillance and prevention and care programs.[20] Thoughtful analysis of effective sampling and recruitment methods are necessary to identify optimal ways to engage BLSGMY in HIV programming to achieve EHE goals. This mixed-methods analysis aimed to evaluate the use of a modified RDS approach and to explore the challenges of traditional RDS assumptions among BLSGMY.

## Methods

This analysis was conducted using baseline RDS data that was drawn from the parent intervention study, *Providing Unique Support for Health* (PUSH). The PUSH study utilized a modified recruitment approach which included RDS with targeted seed selection to recruit and enroll eligible BLSGMY who have sex with men to status-dependent randomized clinical trials (RCTs) that compared coach-based support to standard of care for HIV care or prevention.[21] After identifying low propagation of RDS early in the study, we conducted qualitative interviews with a subsample of diverse PUSH participants across the study sites to explore their experiences and challenges in RDS recruitment.

### Setting and participants

*PUSH* was a multi-site study conducted in Baltimore, MD (Johns Hopkins University), Philadelphia, PA (Children’s Hospital of Philadelphia); and Washington, DC (Children’s National Health System and Whitman-Walker Health). In these cities, Black and Latinx populations account for at least half of the population,[22] while BLSGMY are priority populations for HIV prevention. All study sites have substantial clinical and research expertise among SGMY.

To be eligible for enrollment, participants were: aged 15-24; from the three study sites; birth-assigned male; self-identified Black and/or Latinx ethnicity; and reported oral/anal sex with a cisgender man in the prior 12 months. RDS recruits were required to present with a valid RDS referral coupon to the study team. We focused on birth-assigned males to include male identified, trans feminine, and gender variant youth given the sexual and gender diversity and dynamic sexual and gender identities of adolescents.

Qualitative participants were a subsample of PUSH participants, with an effort to obtain a maximum variation sample in terms of number of successful peer referrals. Participants had the option to decline qualitative interviews without any impact on their participation in the parent study.

### Sampling and recruitment

The PUSH study used a modified RDS methodology. This included the use of RDS coupled with targeted recruitment from clinics, physical venues, online including social media, and community outreach. All eligible and participating youth who were direct recruits from these non-RDS sources were then eligible to become RDS seeds themselves and refer other participants. Similar modifications have been implemented in other studies among SGMY.[23]

PUSH seeds and recruits were asked to complete an in-person screening and informed consent. Participants regardless of ultimate enrollment in an RCT were provided with RDS recruitment coupons and asked to return at a later date to obtain secondary recruitment incentives.

Consistent with RDS recommendations and best practices,[24] RDS implementation was informed by formative research conducted among 18 key informants across the three cities.[18] Once PUSH launched, recruitment followed standard RDS procedures[2] with ongoing recruitment monitoring but added the following modifications for youth, based on prior studies.[13] Eligible and participating youth were offered electronic coupons (e-coupons) with which to recruit peers. Seeds and recruiters received a weblink by text message during the study visit. The link directed the participant to a page where they could manage and share e-coupons with peers from their social and sexual networks. Participants could continue to access the weblink after leaving the study visit. Paper coupons and study fliers were also available upon request.

E-coupons took the form of a text message sent to selected peers inviting them to the study and providing a unique numeric code, study telephone number, information on site operating hours, and the e-coupon expiration date. Peers were asked to display the unique e-coupon code at screening. Text messages contained no information that the study was specific to BLSGMY populations nor focused on the topic of HIV, though participants were encouraged to discuss this verbally with their peers. Initially, participants were offered up to 5 coupons; however, this was later expanded to 10 coupons, with up to 5 reimbursed. Participants were provided with a $50 incentive for completing the initial study visit activities and $5-15, depending on the site, for each eligible and participating peer recruit.

Qualitative participants were recruited from those who participated in the enrollment visit, regardless of whether they agreed to part participate in one of the RCTs. We aimed to interview approximately 7-12 per city.

### Data collection

Upon enrollment, participants were asked to complete a structured, self-administered survey. The survey included network size questions for RDS weighting. This included questions about the number of SGM who have sex with men that the participant knows, the number of these individuals who identify as Black and/or Latinx, and the number of these individuals who are aged 15-24 in their city. Of these, participants were then asked how many they have seen within the last 30 days.[21]

Qualitative participants met separately with a trained qualitative researcher at their site. Qualitative interviews were conducted in private and follow semi-structured interview guides that explored overarching domains of attitudes, beliefs and experiences with the RDS recruitment strategy with the intent of understanding how to better address any potential challenges associated with inviting their peers/friends to the study.

### Data analysis

Quantitative analysis included descriptive statistics and recruitment diagnostics consistent with RDS, including analysis of RDS recruitment networks, coupon distribution and return rate, and participant self-reported network size. RDSAT and Netdraw software programs were used to create network graphs and Stata Statistical Software, version 15 (College Station, TX) was utilized for other descriptive statistics. Descriptive statistics with frequencies and Chi-square tests were calculated to compare participant characteristics by recruitment source including clinic, venue, and internet-based targeted seed recruitment and RDS recruitment.

Qualitative interviews were audio-recorded, transcribed and de-identified. Transcripts were entered into qualitative data analysis software, NVivo. All transcripts underwent an initial round of thematic analysis led by two trained qualitative analysts. Coding was conducted in tandem for the first three interviews and then codes were reviewed and discussed for consistency across coders and to identify additional codes. Codes were modified until high agreement was achieved between coders (Kappa >0.80). RDS assumptions were used as an analytic framework for thematic analysis as well as general barriers to HIV research among SGM youth. Memos were written for each code. Codes were refined and elaborated during the process of analysis through the constant comparison method. Transcripts were subsequently coded separately and 20% of the overall sample was checked to insure consistency of coding across interviews.

### Human Subjects Considerations

Ethical review was provided by the Johns Hopkins Bloomberg School of Public Health, the University of Pennsylvania, and Children’s National Medical Center Institutional Review Boards. This study received a waiver of parental consent for participants below the age of 18 years. Youth advisory boards were also convened regularly in each city for review and feedback on the parent study methods including RDS approach, study instruments, and the intervention. Transportation via ride-sharing apps and bus tokens were provided to participants with limited transportation to minimize research disparities associated with transportation barriers.

## Results

### Quantitative Results

Between August 2017 and October 2019, a total of 442 participants were recruited and 405 enrolled into the study, including 305 (69.6%) seeds. A total of 1,670 coupons were distributed with 133 returned, yielding a 0.079 return rate. RDS networks remained small, with 4 waves being the maximum recruitment depth among seeds that propagated (Figure 1). In terms of network size, participants reported knowing a median of 8 (IQR: 3-20) SGM individuals. This reduced to a median of 5 (IQR:2-15) when asked about Black or Latinx SGM and remained at a median of 5 (IQR 2-10) when asked about those who were aged 15-24 and living in the three cities. Ultimately, participants reported having seen a median of 3 (IQR 1-5) of these peers within the past 30 days. Thirty-eight (9.5%) of participants reported knowing no other peer who identified within this population. Participants requested a median of 5 coupons (IQR: 3-5) for recruitment.

**Figure 1.**
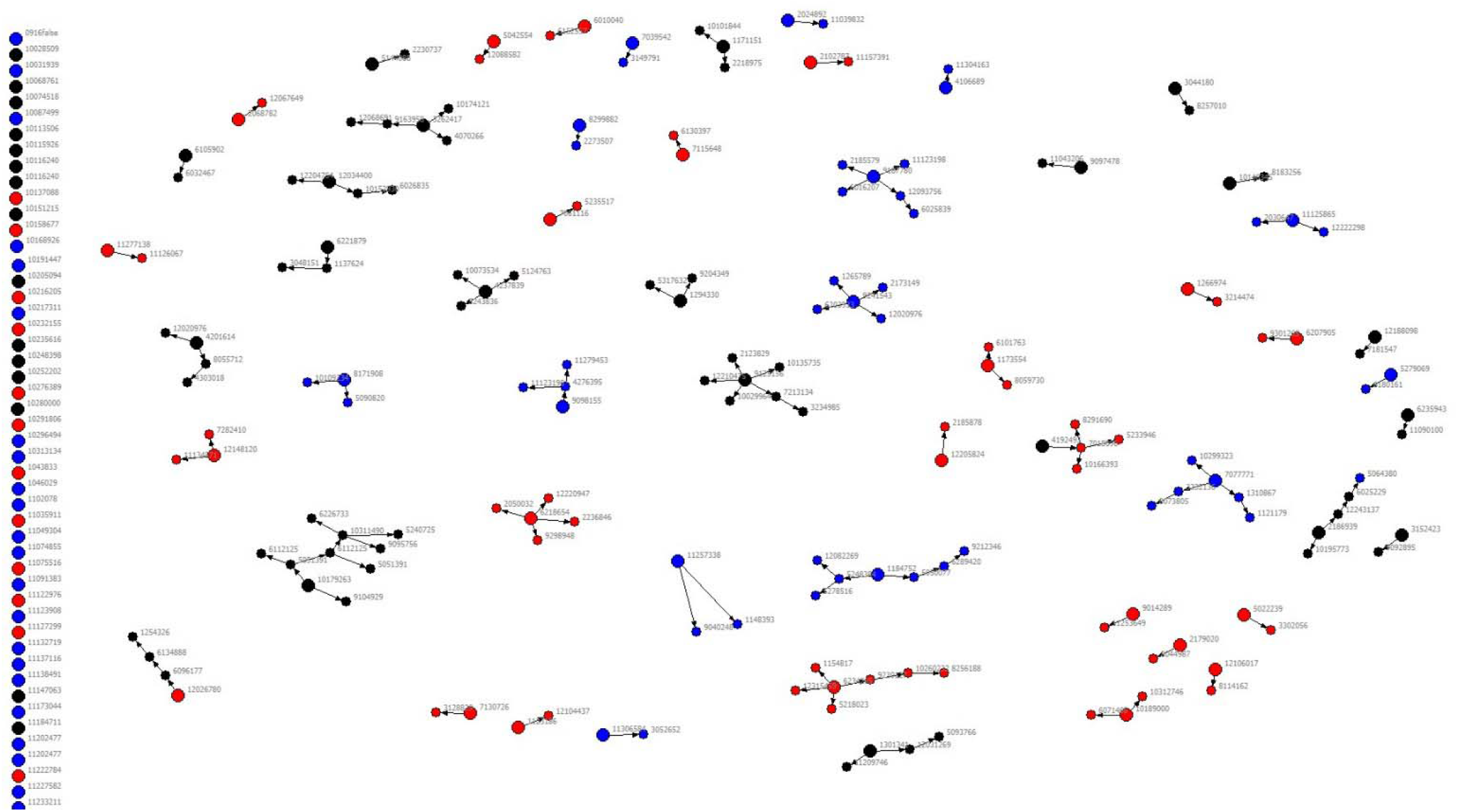
RDS network recruitment diagram: recruitment of Black and Latinx SGMY in Baltimore, Washington DC, and Philadelphia. Legend: Red: Baltimore; Blue: Washington, DC; Black: Philadelphia; Large nodes represent seeds

Clinic-based targeted recruitment produced the largest subsample with 168 enrolled participants (41.5%), followed by RDS (n=123, 30.4%) and substantially lower among venue (n=77, 19.0%) and internet-based targeted seed recruits (n=37; 9.1%). There were few observable differences between participants recruited via these sources. Table 1 describes characteristics of study participants stratified by their recruitment source as RDS recruits or seed recruits. Targeted seed recruitment from clinic-based settings were more likely to result in enrollment of participants who were less likely to report unstable housing in the last year, more likely report a prior positive HIV diagnosis, or more likely to report PrEP use compared to other sources of recruitment. Participants recruited through targeted seed recruitment via the internet were more likely to report Latinx ethnicity compared to other sources, though the sample from this source was limited.

**Table 1.**
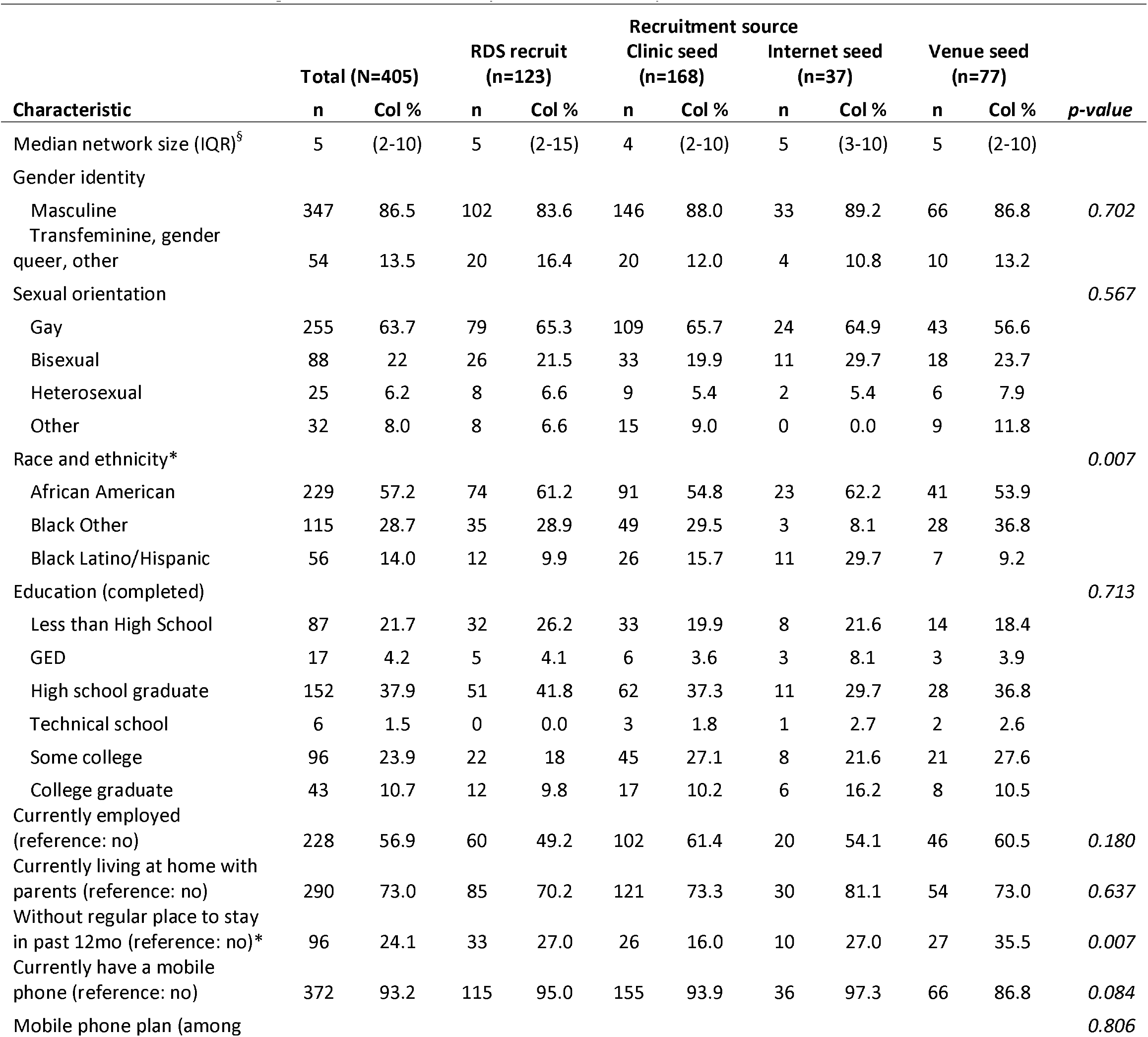
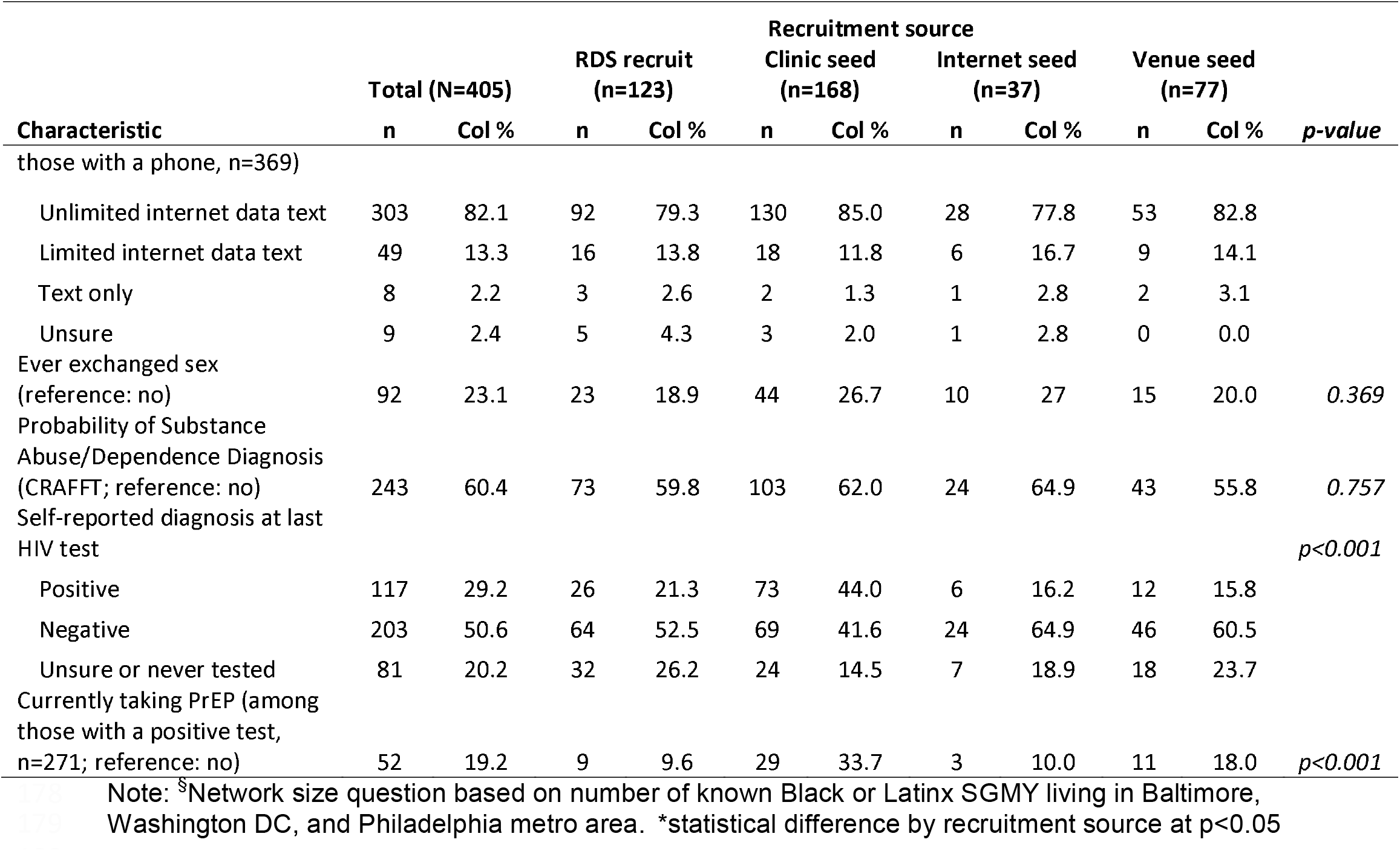
Demographic and other characteristics of Black and Latinx SGMY participants in Baltimore, Washington DC, and Philadelphia, stratified by recruitment source.

### Qualitative Results

A total of 27 youth participated in in-depth interviews between May 2018 and December 2019, including gay or bisexual cisgender men (n=23) and transgender or gender variant youth (n=4). Table 2 displays the demographic characteristics of qualitative participants from the three sites. Most qualitative participants reported difficulties in recruiting Black and/or Latinx SGMY peers, which challenged the success of RDS. Several of the challenges reported by qualitative participants directly affect the assumptions underlying RDS.

**Table 2.** Description of Black and Latinx SGMY participants of qualitive interviews (N=27)

| Site | ID | Gender | Age | Coupons Requested | Coupons Distributed | Coupons Redeemed | Coupon Return Rate |
| --- | --- | --- | --- | --- | --- | --- | --- |
| Baltimore | 1 | Cisgender man | 23 | 5 | 4 | 4 | 100% |
| Baltimore | 2 | Cisgender man | 22 | 5 | 3 | 3 | 100% |
| Baltimore | 3 | Cisgender man | 20 | 5 | 0 | 0 | x |
| Baltimore | 4 | Cisgender man | 23 | 4 | 3 | 0 | 0% |
| Baltimore | 5 | Transgender woman | 24 | 5 | 2 | 2 | 100% |
| Baltimore | 6 | Cisgender man | 16 | 5 | 5 | 0 | 0% |
| Baltimore | 7 | Cisgender man | 17 | 3 | 1 | 1 | 100% |
| Baltimore | 8 | Cisgender man | 19 | 5 | 2 | 2 | 100% |
| Baltimore | 9 | Cisgender man | 21 | 1 | 0 | 0 | x |
| Baltimore | 10 | Cisgender man | 23 | 3 | 2 | 2 | 100% |
| Baltimore | 11 | Cisgender man | 17 | 1 | 0 | 0 | x |
| Baltimore | 12 | Cisgender man | 19 | 5 | 5 | 3 | 60% |
| Philadelphia | 1 | Cisgender man | 21 | 7 | 5 | 2 | 40% |
| Philadelphia | 2 | Cisgender man | 22 | 5 | 4 | 2 | 50% |
| Philadelphia | 3 | Cisgender man | 21 | 9 | 5 | 5 | 100% |
| Philadelphia | 4 | Transgender woman | 21 | 5 | 0 | 0 | x |
| Philadelphia | 5 | Cisgender man | 17 | 5 | 3 | 3 | 100% |
| Philadelphia | 6 | Cisgender man | 16 | 5 | 4 | 4 | 100% |
| Philadelphia | 7 | Transgender woman | 17 | 5 | 0 | 0 | x |
| Washington, DC | 1 | Cisgender man | 18 | 3 | 2 | 2 | 100% |
| Washington, DC | 2 | Cisgender man | 18 | 3 | 1 | 0 | 0% |
| Washington, DC | 3 | Cisgender man | 18 | 3 | 1 | 0 | 0% |
| Washington, DC | 4 | Cisgender man | 23 | 5 | 4 | 0 | 0% |
| Washington, DC | 5 | Transgender woman | 19 | 3 | 0 | 0 | x |
| Washington, DC | 6 | Cisgender man | 19 | 5 | 0 | 0 | x |
| Washington, DC | 7 | Cisgender man | 20 | 1 | 0 | 0 | x |
| Washington, DC | 8 | Cisgender man | 20 | 7 | 4 | 4 | 100% |

### RDS Assumption 1: A networked population

RDS requires a population to be well-networked for the sampling process to function appropriately. One of the most salient themes across all interviews was the reported low connectivity across the population of BLSGMY and very small, tightly knit networks (Table 3; Online Appendix 1). Reasons for a small number of peers outlined by participants included mistrust among peers and simply not knowing many peers that identified as gay or bisexual men, transfeminine, or gender variant. Participants also struggled to identify peers that met the eligibility requirements for age and race. Peers were frequently described as older than the 15-24 year eligibility requirements and/or were not Black or Latinx race/ethnicity.

**Table 3.** Social contexts of Black and Latinx SGMY and relationship to RDS assumptions of networks and reciprocity: explanatory quotes from qualitative participants.

| DOMAIN | CONSIDERATION | EXPLANATORY QUOTE |
| --- | --- | --- |
| NETWORK |  |  |
|  | General small social network | "I would, like I think I said earlier, I don't really interact with too many people day-to-day, a lot of time I spend at work or with son- so If I did have paper ones [coupons], it would be a better |
|  |  | option, I would prefer to do it that way but like I said my access to people is sort of limited.” – *Baltimore 4*<br>“I think... well, for me, my biggest challenge was knowing people that I would want to send it to, let alone would actually do it, but I feel like, you know, other people have more friends than I do. So I don't think that would be a problem for everyone...” – *Washington, DC 4* |
|  | Small number of peers that fit sexual orientation eligibility criteria | “I know millions of females that I could have gave this too, you know what I mean, instead of just men. I don't really know that boys. Gay boys don't hang with gay boys that much.” – *Baltimore 3* |
|  | Small number of peers that fit the age or race/ethnicity eligibility criteria | “Yeah. People that was black and Latino. I don't have too much friends that are black and Latino. They're all white. And I have some black friends.” – *Baltimore 6*<br>“Well, I couldn't bring any friends in. I tried. It was just there are-- I'm the youngest in the house. I'm 21. And all the other girls in the house is, like, 27, 26, and 30. So, they wasn't able to make it in.” – *Philadelphia 4* |
| RECIPROCITY |  |  |
|  | Referring strangers to the study | “I don't know. If it was random, then I probably would be like “No.” I don't know. I would just ask for proof first and making sure that it's not something out of the ordinary, something crazy or something like that. [Interviewer: So you feel like it works better if somebody from your circle asks you to do it.] Yeah, people that you know, it would get them to come in easier without the whole being scared to do it.” – *Philadelphia 1* |
|  | Use of social media | “Facebook. I asked people on the social apps I'd be on, whether if it's Jack'd or Grindr, asked them if they wanted to come in, or I sent them certain information. Some of them have said yes, that they would, but didn't work out too well. [Interviewer: So posting on social media, do you think that that has worked?] Even for a response, people have responded, but I would never- well, I would, but I can never just walk up to somebody and be like ‘Oh, guess what? This and that,’ because I don't know whether or not that would be appropriate or not.” – *Baltimore 2* |

### RDS Assumption 2: A relationship is reciprocal

RDS assumes that peer relationships are reciprocal (undirected). Frequently this is understood that *Peer A* knows *Peer B* sufficiently well to recruit *Peer B*, but also that *Peer B* knows *Peer A* sufficiently well to recruit *Peer A* – i.e. they are not strangers to each other. This assumption appeared to be less frequently violated (Table 3; Online Appendix 1). The majority of respondents expressed that that they would be skeptical if approached by a stranger and reported preferentially referring peers they trusted. Participants also indicated that research study recruitment was not a priority when conversing with acquaintances or strangers.

Though participants were not asked to recruit via social media or dating apps, two participants reported using social media applications (Facebook, Jack’d and Grindr) to recruit individuals. One participant expressed more comfort recruiting strangers over social media platforms than in person. Another participant described building rapport with strangers via social media and then providing study information, which successfully supported peer referral.

### RDS Assumption 3: Recruitment of each individual is random

An inherent assumption underlying RDS is that individuals randomly recruit from within their network. Participants, however, frequently reported seeking to recruit peers who they anticipated would participate in research (Table 4; Online Appendix 2). This was characterized as targeting peers for recruitment who they perceived could benefit from study participation or in need of material resources. Financial incentive was frequently reported as a driver for individual participation. Conversely, participants reported avoiding referring peers that may have difficulty completing surveys due to literacy constraints, low perceived likelihood of participating, or who were past sexual partners.

**Table 4.** Social contexts of Black and Latinx SGMY and relationship to RDS assumptions of random recruitment and sampling with replacement: explanatory quotes from qualitative participants.

| DOMAIN | CONSIDERATION | EXPLANATORY QUOTE |
| --- | --- | --- |
| RANDOM RECRUITMENT |  |  |
| | Characteristics of peers targeted for recruitment | “Sorry, I asked those five people just because I knew that they would be interested in giving their input and basically the research. I wouldn’t ask any other random people because they probably wouldn’t be as interested, but I knew people that I hang out with, people who I know who do outside work in the community would be interested in wanting to work with the research.” - *Philadelphia 2*<br>“Because y’all got to understand, a lot of youth are homeless and what-you-call-them, so a lot of times food vouchers or food things and money is definitely going to-- will wheel a youth in. That’s how I started, struggling. ‘This is a little \$30-\$40 survey,’ boom. ‘They got food vouchers, too, and you bring this,’ dah, dah, dah. Yeah, youth struggle so you never know what the struggle might be” – *Baltimore 5*<br>“[Interviewer: Are there certain kinds of people that you feel more comfortable for inviting?] Or somebody that already needs to get tested. Like you always need to get tested, so why not join the study where you can benefit from it and still get tested and still help other people?”<br>– *Washington, DC 5* |
|  | Characteristics of peers avoided | “I feel like some of my friends don't know how to read or spell. I don't know... I don't know |
|  | for recruitment | if they would be able to get through it [the survey].” – *Baltimore 6*<br>“That's another thing that's holding me back is that a lot of these people that I would send it to, past hookups, I do not... I don't even want them in my phone really, so I don't even want to look through them because all of them were unsafe sexual encounters and so they would not even, you know, look at something like this... I don't think that they even would want to come into Whitman-Walker, you know, so. But they're the ones who need it the most, so.” – *Washington, DC 4* |
|  | Comfort and benefits associated recruiting close friends | “Yeah. I know more so because I was limited to the number of people I could refer, I sought out my close friends more so than other people that I just knew that would've been qualified for the survey, because I wanted to let them know about the opportunity more so than someone that I barely knew.” — *Washington, DC 1*<br>“Yeah, there's still people that I could've invited that I'm like.. "Uh-uh, I don't know," just because like I was just saying, I don't know what their situation is right now and I don't want something to pop up on their phone that they don't want. Also I just don't, I don't know, I still feel like it can get back to me for some reason. <laughs> That's a hesitation that I had.” – *Washington, DC 4* |
| **SAMPLE IS SELECTED WITH REPLACEMENT** |  |  |
|  | Challenges associated with research practice of sampling without replacement | “A lot of names had came to mind, but then the person that recommended me also recommended them because our friend groups, they overlap. So then it was like, "Oh, they already did the study," so then I couldn't invite them.” — *Washington, DC 3* |

Trust was critical to feeling comfortable sharing study information with peers. Participants preferentially referred peers who they felt could be trusted to keep their involvement confidential. The idea of participating in the study with friends was also reassuring and appealing to some participants. Further, participants described having a fear of unintentional peer disclosure associated with the recruitment process and preferentially disseminated coupons to close friends who already knew about their sexuality.

### RDS Assumption 4: Sample is selected with replacement

RDS, particularly the analytic estimators, assumes that the sample is selected with replacement. This assumption is frequently violated by study designs that prioritize sampling without replacement and exclude recruits who have previously participated in the research. The design of sampling without replacement also challenged recruitment for participants with small dense networks (Table 4; Online Appendix 2). Recruitment opportunities were limited for participants whose entire peer network had already participated in the study. Participants reported feeling that recruitment was a competition due to overlapping networks.

### Barriers to Engaging in Research

Challenges to RDS assumptions existed against a backdrop of competing priorities and situational barriers that broadly challenged recruitment of BLSGMY to HIV-related research. Competing priorities for youth, such as work, school and their health were priority over peer referral to a study (Table 5; Online Appendix 3). Participants shared that concerns about drug testing, fear of needles and concerns of breached privacy associated research participation posed challenges in peer referral and participation.

**Table 5.** Barriers to engaging in HIV research among Black and Latinx SGMY: explanatory quotes from qualitative participants.

| CONSIDERATION | EXPLANATORY QUOTE |
| --- | --- |
| Competing priorities | "No, I didn't think about not inviting people, but it's like I didn't think about that, like thinking about, oh this- you know, like just going out here, like, 'You should come to PUSH.' I wasn't thinking about that. I don't know, that wasn't on my mind. I'm more thinking about what's going on with HIV and school and stuff like that. I wasn't thinking about coming right back." – *Baltimore 11* |
| Fear/Skepticism in research participation | "I mean, it was kind of, like I said before, a little nervous because I didn't know what exactly all the ins and outs of the research and what was it geared to. They just told me 'We're just trying to find information to better the community,' but I'm like 'Better the community how? There's so many things that can be worked on or can be addressed,' and then I was just a little bit nervous asking or giving my input on things that I've gone through in my life that I probably wouldn't share with any other body, but being as though it's research, you need to get all those variables of everything so you can have data or whatever. But you know, at first I was like I would like to do it just because I want to make a difference and I want my input to be in the research but having those feelings like what questions or what I have to answer or what you guys want to know was in the back of my mind." – *Philadelphia 2*<br>"[Interviewer: Do you ever feel apprehensive about inviting people to join the study?] Sometimes, because I don't want them to question. Like, 'How the hell do you know about this?' Yeah, people are very nosy and they just... – *Washington, DC 5* |
| Situational Barriers | "A lot of people don't have IDs, and it was a requirement for you to have your ID. That's the only thing that was a problem with me. There's a lot of-- I know a lot of my friends really don't have IDs. They got warrants and stuff, so they're not trying to go up there and get it. But I understand why ya'll ask for ID, too, so y'all can be sure it's legitimate. But yeah, the ID part." – *Baltimore 5*<br>"I told-- I brought in, I only brought in three [peers]. But I told, probably at the most, likely 10 people. I just couldn't make it. That's another concern. People can't make it so if they had transportation to get here it would work out. [Interviewer: Okay, so you think transportation was an issue?] With-- For most of them. [Interviewer: Okay, so the people that came into the study, those three people, what do you think made it easier for you to invite them and for them to come in?] They live close by." – *Baltimore 12*<br>"[Interviewer: .. talk a little bit about how the text and your phone being broken was a challenge?] Yeah, so when I had lost my phone, it was hard to even remember about the coupons because I didn't have that reminder in my face. I forgot about the coupons." – *Washington, DC 5* |
| Sexual Orientation | "Challenges? I'd say one challenge would be not being out of the closet but generally, generally speaking, if you go around an organization, if you participate in the study, you're most likely out of the closet. I mean, other cases, they're really not. So, I think in that case maybe people are scared that if they hand this out, then someone is going to know that they're gay or somebody down the line can tell someone that this person gave them this and they want to take their time to come out and make sure their parents or whoever are hearing from them and not someone else...." – *Philadelphia 3* |
|  | "I would say one main thing is like, say if you were to do the survey in secret and say you're not fully comfortable in your identity, then to pass out the coupons to whoever would in certain kind of imply something about yourself. So I feel like for those who aren't necessarily firm in who they are and their identity yet, then that would be one reason that they don't pass them out...." – *Washington, DC 1*<br>"[Interviewer: ... Do you have any recommendations for how we can improve that coupon process, either digital or paper?] I don't know. I feel like for the black and Latino community, like privacy when it comes to matters of sexuality is pretty big, so just like keep them interesting. Like your information is going to be private. Like as long as you don't tell anyone, no one's going to find out you're participating, and that whatever their HIV status is, no one's going to find out unless they're sharing it themselves. ... Yeah, just privacy, privacy, privacy." – *Washington, DC 8* |
| HIV Status | "Some people are because some people are actually scared to know their results. Like me for instance, I was young when I found out everything, so yeah. I can't lie. Now, I really would be scared to get my results because at this point in time, I'm a escort and everything, so me dealing with so many people and sexually-wise and stuff, I would really be scared. I'd be like, 'Girl, I don't need this' or something like that." – *Baltimore 5*<br>"...But also just I think feeling like they're giving some confidentiality away if they recruit other people or maybe they're worried will I run into them, you know, coming here. Yeah, I think there's just.. not a stigma but like when you hear.. if someone you know... if I told somebody 'I'm doing a study at Whitman-Walker,' they'd be like, you know, 'Why there, do you have HIV' you know, 'What's going on.' So I think people want you know, to just keep that to themselves maybe." – *Washington, DC 4*<br>"[Interviewer: And I also wonder how that went, if you told anybody that there was HIV testing with the study?] That part I didn't mention. And, like, at the one place I didn't know that I had to in order to receive a gift card. And I don't know. Like, it's not a bad thing, it's just suspicious. For me, it's a little scary, because I'm private about it... Like, I don't like too many people knowing or-- because it's my business. It's like I'm a private person. I don't want everybody, like, to know, 'Oh, she has HIV.' Because people are so judgmental and they'll always say, 'Oh, well, you know, she does this, that, and a third. So, she has HIV.' People are rude. So, I just have to play my cards right." – *Philadelphia 4* |

Situational barriers also posed challenges for peer recruitment. Participants described facing personal challenges in recruiting peers that prevented them from completely participating in RDS. For example, some participants lacked or had inconsistent access to a cell phone, thus limiting the sharing of study information. Participants also reported lacking transportation that challenged study visits and some reported that peers did not possess a government-issued form of identification that was required for study enrollment.

Concerns related to privacy and confidentiality impacted not only willingness to recruit peers but also individual willingness to participate in the study. Participants who had not disclosed their sexual orientation or HIV status described a reluctance to participate in the study due to concerns of privacy and confidentiality (Table 5; Online Appendix 3). The focus of the study also mattered. Multiple participants expressed apprehension about being HIV tested or involved with an HIV focused study due to concerns of unintentional disclosure of one’s HIV status, behavioral risks for HIV, or perceptions of one’s HIV status.

## Discussion

Use of RDS to sample BLSGMY faced challenges in this multisite study in the US. RDS seeds and recruitment networks failed to propagate beyond a limited number of waves, despite participant reports of median network sizes of approximately 5 peers. While a network size of 5 peers seems feasible for peer referral, other research has shown that SGMY were 60% more likely to effectively refer at least one peer when they had a network size of 10 or more.[13] Coupling RDS with targeted recruitment from physical and online sites, however, supported access to and engagement of over 400 BLSGMY. A similar modified approach (“Starfish Sampling”) was recently reported by Raymond and colleagues for sampling transgender men in San Francisco, which permitted participants who were recruited via venue-based sampling to then refer peers using standard RDS methods.[23] While limited in its ability to produce population estimates, the authors noted that “starfish sampling could be considered for recruitment in populations when available tools are inadequate”.[23]

Our qualitative data and that of others,[13-15] however, suggested that BLSGMY infrequently have the well-developed networks that are observed among adults. Youth reported knowing and associating with peers who identify with the same race/ethnicity or who identify as SGMY but may not have peers who identify across these characteristics. Under-developed peer networks may be due in part to emerging sexual orientation and gender identities, and limited disclosure to peers of the same age range or race/ethnicity. Further, BLSGMY are more likely to experience marginalization, stigma, and isolation, which may make unequally compel some youth engage in sampling strategies, while turning others away.[18] Targeting sampling to groups on the basis of race/ethnicity, age, gender identity, and sexual orientation as well as common research practices of sampling without replacement may inherently break underlying social networks and violate the network assumption that is fundamental to RDS. RDS and related modified network-based approaches among BLSGMY that are less restrictive in eligibility criteria or permit inclusion of other peer populations, such as female members of social networks, can potentially help to bridge networks and promote engagement in research.[25]

Other RDS assumptions that are critical to implementation and analysis were also reportedly violated by youth participants. Youth frequently reported preferential recruitment of peers who either needed a financial incentive or would be interested in research. While well-intentioned, these practices may introduce bias and, in the case of recruiting to status-dependent RCTs, may challenge engagement of those who are not engaged in but could benefit from HIV prevention and care. This is also an important consideration for studies that select RDS for its reported ability to estimate population means, as preferential recruitment has been associated with biases in estimation.[26]

The future of RDS among BLSGMY in the US may rely on greater adaptation to technology but must do so with consideration for RDS assumptions, particularly if used to generate population-based estimates. Some participants described use of social media and dating apps to distribute e-coupons, suggesting this is a viable option; however, it may also violate assumptions of reciprocity depending on the nature of the relationship within social media. Research has shown that youth view social media friends and followers as sources of social support;[27, 28] thus, youth may experience similarly strong or stronger emotional ties with online peers as they do with peers they regularly see in-person. Social media has recently been integrated into RDS approaches among SGMY, improving enrollment despite that unique race and socio-economic differences were observed when compared to traditional RDS and to nationally representative samples.[14, 29] The authors of a webRDS study acknowledged racial disparities in consistency of computer and internet access,[29] drawing important consideration to the possibility for webRDS to potentiate disparities in health research. Taken together, these studies highlight the potential of social media to diversify samples recruited through RDS, but also suggest that sampling methods using social media alone may miss important populations who could benefit from public health interventions.

Study findings should be viewed in light of limitations. First, the proportion recruited via RDS may be underestimated, as anecdotal reports from staff suggest that coupons were provided to peers who participated in the study, but who forgot to display the study coupon and were possibly misclassified as targeted seeds. Finally, the sample of transgender and gender variant youth enrolled in the qualitative and quantitative components of this study are small and unlikely to be representative. Other studies have recently faced similar challenges with the use of RDS to sample these populations, highlighting the importance of identifying a sampling method that is acceptable to transgender youth.[14, 23]

## Conclusions

Traditional RDS may have a limited role in sampling SGMY, particularly those who are racial or ethnic minoritized populations in the US but may be improved through coupling with other sampling approaches and/or integration with social media platforms. Sexual and gender identity formation and peer disclosure are evolving processes among BLSGMY, potentially resulting in small social networks with varying degrees of trust and challenging traditional RDS assumptions. Research hesitancy and situational barriers also present barriers to individual recruitment and peer referral that may be addressed in study design.

## Data Availability

Data available upon request.

## Acknowledgements

This study was supported by the National Institutes of Health (R01DA043089) through the NIDA and the Johns Hopkins University Center for AIDS Research (P30AI094189). The authors acknowledge the contribution of the investigators and staff of PUSH Study Group at participating research sites, including the Johns Hopkins University, Baltimore, MD (F Shorrock, J Conley, and A Alvarenga); Children’s Hospital of Philadelphia, PA (M Castillo, N Dowshen, A Schlupp, A Lopez, and W Vickroy); Children’s National Medical Center and Whitman Walker Health, Washington, DC (C Trexler, LJ D’Angelo, J Kwait, R Carr, J Leslie, and B Smith). The authors also thank the Johns Hopkins University and Children’s Hospital of Philadelphia’s Adolescent Medicine Teen Advisory Boards, who provided feedback on the study materials. The authors thank all the adolescents and young adults who participated in this project for their time and effort.

## Author contributions

RS, DC, CB, NG and ALW designed the parent study; RS and DC are the joint principal investigators. KHF and DB conducted the interviews, JI and a research assistant conducted the qualitative coding with oversight by RS. ALW and IJ jointly reviewed and analyzed the data. ALW led the write up of the findings with contribution from IJ. All authors have reviewed and provided input to the manuscript.

