## Supplementary materials for "An evaluation of assumptions underlying respondent-driven sampling and the social contexts of sexual and gender minority youth participating in HIV clinical trials in the United States"

**Supplemental Materials**

**Appendix 1.** Social contexts of Black and Latinx SGMY and relationship to four RDS assumptions: explanatory quotes from qualitative participants

| **NETWORK** |  |  |
| --- | --- | --- |
|  | General small social network | “Real friends is hard to come by nowadays….You can’t trust everybody. Because usually I do not keep friends, because something with me shuts-- suddenly I see one think kind of-- I don’t like that.” -*Baltimore 1* |
|  | Small number of friends that fit the sexual orientation eligibility criteria | “[Interviewer: Were there people that you were less comfortable inviting?] Yes, because I didn’t know how people would take it. But from the beginning, I never really had a lot of options of inviting people, because I didn’t have that many gay friends. So the one I did have, he was ineligible, but the boy I was messing with, he was eligible, and I brung him. But I feel like anyone I’m messing with and they’re out, I’m comfortable with bringing them in. That’s the same thing.” – *Baltimore 7*  “[Interviewer: So as of today have you brought in any peers or friends or people you know to the study?] No, ma'am. [Interviewer: Okay. So can you tell me what did not work for you?] Let me see, the lack of-- how can I say it? The lack of people that I know that's doing the same things I'm doing. I can just say it like that… As far being well, well you know. <whispers> Can I say it? [Interviewer: You don’t have to sign language me. You could say it.] Oh yeah. Well, far as like being like I guess you could say down low, I guess. People don't know about me, you feel me?, I still want to…” – *Baltimore 9*  “[Interviewer: … why might other people not be able to distribute any coupons?] Because like I say, either a lack of not knowing-- by not knowing people that’s in their shoes. Like if they-- people don't know about them, they going to have an issue like around like networking, I’m not going to say networking, but networking to try to find. You know what I'm talking about?” *– Baltimore 9*  “[Interviewer: … is there anything our study team can help you or other people do to distribute coupons?**]** Maybe if there was a, I want to say-- it's hard to-- I was, in my mind, thinking of a resource center because if there is a person who doesn't know any other gay people, then they can't recommend anybody. So if there is places in the D.C. area that aim to help LGBT kids and it's like I would have never, I never knew, known about that. So then, I can go look around, meet people. Then it's, ‘Hey, I did this. Here's a coupon.’ Stuff like that.” – *Washington, DC 3* |
|  | Small number of friends that fit the age or race/ethnicity eligibility criteria | “[Interviewer: But you’ve kind of said there are certain types of people you felt more uncomfortable inviting.] It would only be the people that I’m really, really close to that I’ve known for some years…I don’t know too many people. It’ll just be my family, and they are 30-plus [years], so yeah.” – *Washington, DC 7* |
| **RECIPROCITY** |  |  |
|  | Referring strangers to the study | “[Interviewer: So if any random person comes and tells you about the study you’re not going to?] Mm-mm. Because I’m like ‘Who is you? Why would I go somewhere with you?’ But if it was my best friend or somebody I’m close with or somebody I trust then I’ll probably be, like, ‘Yeah.’” — *Baltimore 11* |
|  | Use of Social Media | “I think they're all strangers actually. I figured- I don't care if I get judged for this. I just downloaded Grindr again since forever and you know how sometimes you get like a free week, or extra.? I was like, oh, I want to take advantage of this. So, what I did was pretty much just used the filters, 18, because it’s 18 on the app.**…** I was like, well, 18 to 24 and I put Latinos. All Latinos and then I messaged all the people on the search bar and then I did the same for the African American community… Pretty much what the [study] website says. I just copy and paste everything [into the message] and then at the end, at the end of saying what the study was about and all that, I would give them a compliment, a true one. I wasn’t being fake about it. I pretty much say ‘Nice tie,’ ‘Cute smile,’ ‘Your hair is gorgeous,’ you know something like that. So, I think that really attracted people. I mean, it’s only four people that came in. I think that’s something. But I think what really attracted people was when I would say ‘Oh, your transportation provided,’ for example. You had to make that clear to people because sometimes people are lazy…So, I think it was more like providing people with the basic information, not just a vague three points on what to do but telling them ‘this is for you. This is for our community. This is for our Latino and for our African American communities, because we need this,’ and people are really connected to that. People really like that.” – *Philadelphia 3* |

**Appendix 2.** Social contexts of Black and Latinx SGMY and relationship to RDS assumptions of random recruitment and sampling with replacement: explanatory quotes from qualitative participants

| **RANDOM RECRUITMENT** |  |  |
| --- | --- | --- |
|  | Characteristics of peers targeted for recruitment | “I was just trying to get everybody I could. I wasn't even worried about asking or being scared. I'm not trying to say I was doing it for the money, but at that time, I didn't have a job. That was the only money I was really getting.” – *Baltimore 6*  “Everybody gonna flip over a fool. I’m gonna tell you the main reason why people are coming: ‘Cause y’all giving them money. That’s the main reason. But me, I need something to do-- money or not, I don’t care. I just need something to do.” – *Philadelphia 6*  “Because you get checked out, and you get to get checked out for free, because any other place-- say if you don’t got no insurance-- mm-hmm…Yeah, if you don’t got no insurance, you going to have to pay, and you have to pay more than the money that you getting paid to come-- mm-hmm.” – *Baltimore 1*  “You’ll get money. Because a lot of young people out here, they like money. Like they money hungry, so if you all say, ‘I’ll give you some money if you bring in friends,’ the first thing they’re gonna say is, ‘How much?’ Then once you tell them, they're like, ‘I got you.’ Because they’re money hungry, especially if you don’t got a job, you will need money. Because I know, my generation, they’ll do- instead of being out here prostituting, they’ll be, like, ‘Well, I got money, I don’t need to go on the stroll tonight. I don’t need to prostitute, I got this, I got that.’ You know what I’m saying? Let’s say you get $15, you bring in 5 friends. That’s, I don’t feel like doing” – *Baltimore 11* |
|  | Characteristics of peers avoided for recruitment | “[Interviewer**:** And what about the people that you just didn’t invite? What made you not want to invite them?] Because I already knew they wasn’t going to come.*– Baltimore 5*  “The people that-- okay, like some of them-- just because people talk a lot, that kind of stuff. Some-- I don't-- I, like I said, I don't know that many people in it like that but, you know, people talk. I don't need all that.” – *Baltimore 9*  “[Interviewer: Were there some friends who-- were there certain types of people who you felt more comfortable inviting or less comfortable inviting?] Certain people I just didn’t ask, because I knew the answer.” – *Philadelphia 5* |
|  | Comfort and benefits associated recruiting close friends | “[Interviewer: So when you talk to people about the study did you talk to certain types of people or were there people you felt more comfortable inviting?] I wouldn't want to say types of people but yeah, I would say types of people. Like, because I told you I don't really know too many people like that, you feel me, that's like that. So it's like the people I do know I was like I know them like personally, so I had no problem coming to them.” –*Baltimore 9*  “[Interviewer: So for the people that you did not invite, can you tell me what may have prevented you from inviting them?**]** We weren’t close. Not as close, but we weren’t talking, and when we were talking, that’s just not something that came up in the conversation. We were barely talking, because if I’m not talking with you for a while, I’m not going to say ‘Hey, by the way—' that’s just something you’re not thinking about.” — *Baltimore 7*  “[Interviewer: Three people? Great. And, sort of I just want you to describe how you actually talked to them.] They were just close friends. So, they heard about me doing it, so they just was like, ‘Oh, let’s just all do it together.’” — *Baltimore 3*  “Because if-- sometimes-- some people have their notifications where like you can read the text. I didn’t want anyone seeing like, you know, “You’re invited to this Whitman-Walker Study,” and then maybe someone knew about Whitman-Walker, and that would kind of out them. You know, you never know.**…**So I try to be very careful with that sort of thing.” – *Washington, DC 8* |
| **SAMPLE IS SELECTED WITH REPLACEMENT** |  |  |
|  | Challenges associated with study practice of sampling without replacement | “Yeah, competition, because what if you bring in them, and after you bring them in, you can’t bring in nobody else, and they’re bringing in all these damn people like-- and they’re just overstocking your budget, your 15 dollars, your little 5, 10, 15, 20, 25, 30 dollars is nothing due to their 10, 20 people who just walked through the door with referrals. So, yeah, it plays a part in competition, too, because me myself, look, if I know that you could do the job better than me and you think that I just struggle my ass off to get this job and you just about to come in here and snatch the job like that, no, I would be highly pissed. I’d be like “Girl, uh-uh, you better find another job. Do not come my way with this job.” So yeah. [Interviewer: It would be frustrating if you invited someone, and then they started inviting people who you could’ve invited. Is that what you mean?] Well, yeah, it’s like-- yes, exactly. ‘Oh, girl, I tried to invite them, and they didn’t come in here, but they in here with you. What’s going on?’ And then extra things open up, and then who has time for that?” — *Washington, DC 7* |

**Appendix 3.** Barriers to engaging in HIV research among Black and Latinx SGMY: explanatory quotes from qualitative participants

| **CONSIDERATION** | ***EXPLANATORY QUOTE*** |
| --- | --- |
| Competing priorities | “I think their [the study] age group has to be higher. If you’re all in your 20s, your age going to have to be higher, because if you’re in high school, your least thing from your mind is ‘Oh, let me hand out—' you see what I’m saying? Because a lot of people in high school are antisocial, so you’re not going to be thinking ‘Oh, let me hand them out’ you know what I’m saying?” – *Baltimore 7* |
| Fear/Skepticism in research participation | “Well, if it's like a drug test and they can easily come in, then it's not nothing bad. I’ll just tell you most of my friends smoke weed and they get scared if they get drug tested. But you're not getting drug tested and you're getting free money. I guess it's worth it.” – *Baltimore 6*  “One of my friends was scared of needles. I told him he might have to get a needle or something. He was scared. So it took him a little while to come in…” – *Baltimore 10*  “I brought two peers into the study. I brought my best friend, and I brought one of my close high school friends when I was in high school. And my best friend was skeptical because of needles. She doesn’t want to get pricked, and she knows a lot about the study. She refuses to get a needle at all. If it’s a swab or something else like a drawing, then sure, but if it’s a needle, she’s most definitely not doing it. My high school friend, she chickened out because of the needles. Majority of my friends that I’ve brought or even referred, they didn’t like it, even because it was all the way up here or they were scared or they were insecure about certain things, or, basically their insecurities.” – *Washington, DC 6* |
| Situational Barriers | “…I’ve really been telling people about it by mouth, because my friends all-- they don’t have a phone…” – *Baltimore 10* |
| Sexual Orientation | "Because either being that person wasn’t cool, we had sexual relations in the past and it was kind of awkward. That’s what maybe it was, because I don’t have a lot of gay friends and such a thing, and I don’t hook up with a lot of gay people, and the ones I do hook up with, they’re DL. They don’t want to hear nothing about that because of, I guess, the fear being seen going in a clinic, and for people who know about this program, there’s always that fear of being exposed and stuff like that, even though it’s 100 percent guaranteed. I just never came to them about it, because it was like we do what we do and we don’t talk on a regular basis, so it’s like-- yeah.” –*Baltimore 7*  “[Interviewer: So as of today have you brought in any peers or friends or people you know to the study?] No, ma'am. [Interviewer: Okay. So can you tell me what did not work for you?] Let me see, the lack of-- how can I say it? The lack of people that I know that's doing the same things I'm doing. I can just say it like that… As far being well, well you know. <whispers> Can I say it? [Interviewer: You don’t have to sign language me. You could say it.] Oh yeah. Well, far as like being like I guess you could say down low, I guess. People don't know about me, you feel me?, I still want to…” – *Baltimore 9* |
| HIV Status | “You’re like, ‘You got to take a urine.’ Yeah, at the consent form. They sign everything and everything. You have to take a urine test and a HIV test and let them know we don’t care about your results. This just is for the study, and it won’t connect your name to it and stuff... There’s a lot who have a problem with the-- it being out there.” – *Baltimore 5*  “Some people might not want to talk to people they don’t know, or they don’t want their friends to know that they’re in the study, or they don’t want people curious about their HIV status. That’s really what it is. You can recruit people just word of mouth, but people are worried about what people gonna think about them. I’m really not that type. ‘Cause I’m good.’” – *Baltimore 10*  “I think their insecurities is because they’re scared that they have it, and they don’t want to face it. My friends are very dramatic …So once you first say something about a needle, it’s like automatic chop; automatically I’m not doing it, or some things that made it hard for them not to-- I don’t know. I think it’s basically they’re scared to face it if they do have it. And I still be talking-- wouldn’t you want to know? Because you don’t want to walk around sick, and then it gets worse and worse all the time, and you’re just sitting there thinking you’re okay and you’re really not. That’s not good at all. That’s just not taking care of yourself as a regular human being. I just think that that’s the-- I swear, I really think that that is the only problem provided I wanted to-- the only two problems: they’re scared to face if they do have it, and the needles.” –*Washington, DC 6* |
